# Mediating Effect of Physical Activity on Relation between Cardio-respiratory Fitness and Physical Function Capacity of Older Adults

**DOI:** 10.1101/2023.06.06.23291026

**Authors:** Eric A. Aloko, Edward W. Ansah, Daniel Apaak, Emmanuel O. Sarpong, Munkaila Seibu, Stephen R. Sorkpor

## Abstract

The ageing process is normally accompanied by several physiological changes like a decline in physical function and increased risk of chronic health conditions. In older adults, maintaining physical function and cardiovascular health is essential for maintaining independence and quality of life. Cardio-respiratory fitness and physical functional capacity (PFC) are two important indicators of physical health in older adults. This study aims to investigate the mediating effect of physical activity (PA) on the relationship between CRF and PFC in older adults. Using quantitative cross-sectional design, we employed a multistage sampling strategy to recruit 998 older adults from Navrongo for this study. The senior fitness test battery, international physical activity questionnaire (IPAQ) short form for elderly, weighing scale and tape measure were used to collect the data. The findings revealed that, 53.4% and 53.9% of these older adults had reduced PFC and CRF respectively. It was also found that PA partially mediates the relationship between CRF and PFC of the older adults with an indirect effect of CRF on PFC as β = .0030, t = 1.579 p < .05, with a direct effect of CRF on PFC, β = .867, t = 42.954, p < .05, and the total effect, β = .872, t = 43.110, p < .05. We concluded that physical activity partially mediates the relationship between CRF and PFC among older adults in Navrongo. Therefore, there is the need for evidenced-based intervention to promote PA among these older adults to improve their PFC and quality of life.

## Introduction

### Background

Ageing is associated with a decline in physical function, which can negatively impact the quality of life of the aged (Nascimento et. al., 2022). According to Forman, Arena, Boxer, Dolansky, Eng, Fleg, and Shen (2017), cardio-respiratory fitness is an essential determinant of physical function in older adults, because it plays a crucial role in the maintenance of activities of daily living (ADLs). In order to maintain physical function in older persons, physical activity is also being recognized as a key element (Langhammer, Bergland, & Rydwik, 2018). Physical function refers to the ability of the individual to perform daily activities and tasks, such as walking, climbing stairs, and lifting objects, without experiencing undue fatigue or discomfort (Denteneer, Van Daele, Truijen, De Hertogh, Meirte, & Stassijns, 2018). Additionally, the ability to maintain physical function is essential for independent living, quality of life, and prevention of disability in older adults. Myers, Kokkinos, Arena and LaMonte (2021) espoused that, PA and cardio-respiratory fitness are important modifiable factors that contribute to physical function in older adults. Physical activity is any bodily movement produced by skeletal muscles that results in energy expenditure (World Health Organization, 2010), whereas cardio-respiratory function refers to the ability of the cardiovascular and respiratory systems to supply oxygen to the muscles during daily physical activity (Wu, 2022).

Several studies have demonstrated that physical activity is positively associated with physical function in older adults (Sales, Levinger, & Polman, 2017; Layne et al., 2017: Dugan, Gabriel, Lange-Maia, & Karvonen-Gutierrez, 2018). According to O’Neill, and Forman (2020), physical activity improves muscle strength, balance, and endurance, all of which are important determinants of physical function. In addition, physical activity reduces the risk of chronic conditions, such as cardiovascular diseases, obesity, and diabetes, which in turn negatively impacts physical function in older adults. Cardio-respiratory function is a significant determinant of physical function in older adults (Kwok, So, Heywood, Lai, & Ng, 2022). Furthermore, cardio-respiratory function is positively associated with muscle strength, balance, and endurance, and has been shown to decline with age (Liguori et al., 2018) Additionally, low levels of cardio-respiratory function is associated with an increased risk of disability, chronic conditions, and increase mortality in older adults. The combined effect of physical activity and cardio-respiratory function on physical function has been studied extensively, with evidence suggesting that these factors have independent and additive effects on physical function capacity in older adults (Franklin, Wedig, Sallis, Lavie, & Elmer, 2023). However, the underlying mechanisms through which physical activity and cardio-respiratory function influence physical function are still not well understood (Omar, Pandey, Haykowsky, Berry, & Lavie, 2018).

It is estimated that Ghana would see a rapid increase in the ageing population, associated with a rise in non-communicable diseases (Bawah et al., 2016). Therefore, understanding the relationship between cardio-respiratory function, physical activity and physical function ability of older adults is essential for the development of appropriate interventions to promote healthy ageing. One potential mechanism through which physical activity may influence physical function is by mediating the relationship between cardio-respiratory function and physical function (Marques, Mota, Viana, Tuna, & Figueiredo, 2012). Despite the potential importance of physical activity as a mediator in cardio-respiratory function and physical function relation, few studies have explored such among older adults (Omar et al., 2018). Therefore, this study aims to investigate the mediating effect of physical activity on the relationship between cardio-respiratory function and physical functional capacity in older adults in Navrongo. It is hypothesized that, physical activity participation mediates the relationship between cardio-respiratory function and physical functional capacity in older adults residing in Navrongo. The findings may provide insights into the potential mechanisms through which cardio-respiratory function and physical activity contribute to the maintenance of physical function capacity in older adults in Navrongo and inform the development of evidenced-based interventions to promote healthy ageing in this population.

## Materials and Methods

This study employed quantitative cross-sectional design to investigate the mediating effect of physical activity on the relationship between cardio-respiratory function and physical function capacity of older adults in Navrongo. One advantage of this design is that it is useful for providing a broad understanding of a population’s characteristics, attitudes, or behaviors. It is also relatively easy and cost-effective to conduct, making it an attractive option for researchers (Wang, & Cheng, 2020).

We sampled 998 senior citizens, representing 60% of the total population (1663) from 1142 household, using a multistage sampling strategy. First of all, the stratified sampling technique was employed to put the entire population into two strata (male and female). The quota sampling was then employed to sample 60% from the male population (722) and 60% from the female population (941), given a sample of 433 males and 565 females. According to Vasileiou, Barnett, Thorpe, and Young, (2018), 60% and above sample size of a population is good representation of the entire population. In an instance where time is of essence and limited availability of resources, 60% of the population was ideal to generalize the findings of the sample to the larger group. According to Mafuure (2017), the quota sampling is a simple yet effective way to do research. With the quota sampling, the researcher must put the sample into mutually exclusive sub-groups then a quota is calculated from each sub-group. The merits of this sampling procedure are that, it is good if the researcher is working within a limited time period. Additionally, it enhances the representation of any certain subgroup within the population, preventing these groupings from being over represented. (Sharma, 2017). However, the quota sampling may be prone to bias as the researcher is likely to assign a greater percentage to a particular group. But in other to be able to make proper comparison of the various sub-groups, the researcher will assign equal percentages to all sub-groups so that all groups will be equally represented. Navrongo is already grouped into four cardinal zones (North, South, East and West). The North, South, East and West comprises 7, 9, 7 and 6 suburbs respectively. The sample size for each stratum (433 males and 565 females) was divided into four and assigned to each zone making it 108 males and 141 females each for the following zones (North, East and West) except for South which comprised 109 males and 142 females. The suburbs from each zone were labelled S1-S7 for North zone, S1-S9 for South zone, S1-S7 for East zone and S1-S6 for West zone. The North and East with 7 suburbs each will be assigned 15 and 16 males each for the first 4 (S1-S4) and last 3 (S5-S7) suburbs respectively. Also, 20 and 21 females each came from the first 6 (S1-S6) and 7^th^ (S7) suburb respectively. For the West zone with 6 suburbs, each suburb (S1-S6) was assigned 20 males, while 23 and 24 females each came from the first 3 (S1-S3) and last 3 (S4-S6) suburbs respectively. Finally, the North zone with 9 suburbs comprised 12 and 13 males each for first 8 (S1-S8) and last 1 (S9) suburb respectively. While, 15 and 16 females each came from the first 2 (S1-S2) and the final 7 (S3-S7) suburbs respectively.

Five research assistants were assigned to each zone. Since there was no available data on the number of older persons in the various households, the research assistance visited the various suburbs and houses in each zone to conveniently sample as many as were available until the required number of males and females for each suburb was attained to give a total sample of 433 males and 565 females.

### Setting

The Kasena Nankana Municipal is one of Ghana’s 260 Metropolitan, Municipal and District Assemblies (MMDAs) and is a member of the Upper East Region’s fifteen (15) Municipalities and Districts, with Navrongo serving as the regional administrative center. Within the Guinea Savannah woods is Navrongo. Between latitudes 11o10’ and 10o3’ North and longitude 10o1’ West is about where it is situated. The town shares boundaries with Paga and Bolgatanga, Sandema. However, it is also bounded to the north of the Ghana by Burkina Faso. The number of household in Navrongo stood at 32,000 with a total population 109, 944 with 53,676 males and 56,268 females (Ghana Statistical Service, 2013). The main occupation of the people in the Municipality is farming and trading. The built environment of the Municipality is not exercise friendly. Also, there exist a poor culture towards meaningful physical activity and exercise in the Municipality. Hence, sedentary living is gradually becoming an order of the day in the Kassena Nankana Municipality.

### Instruments

We used instruments adopted from four preexisting instruments, namely, the Senior Fitness (Functional Fitness Test) battery, Omron weighing scale, tape measure, and international physical activity questionnaire (IPAQ) Elderly short form. Rikli and Jones created the Senor Fitness Test battery as part of Fullerton University’s lifelong wellness programme. The test is frequently referred to as the Fullerton Functional Test as a result. It is a straightforward, user-friendly battery of tests that evaluates older individuals’ functional fitness. The test is easy to understand and effectively test the aerobic fitness, strength and flexibility among adults, using minimal and inexpensive equipment. Within the test battery are specific tests and their reliability coefficients; “chair stand test (.88), arm curl test (.79), sit and reach test (.81), back scratch test (.82), up and go test (.84) and 2-minute step up test (.81)” and a composite reliability of .83 for PFC (Rikli & Jones, 2013). Reliability coefficients estimated based on the current study are .79, .80, .77, .78, .80, .79 for chair stand test, arm curl test, sit and reach test, back scratch test, up and go test and 2-minute step up test respectively.

The IPAQ-Elderly short form was used to measure the physical activity levels of older adults in Navrongo. This instrument was tested in six centers involving 12 countries across all continents and a reliability and validity values of .81 and .84 were reported under Africa (Bădicu, 2018; Craig et al., 2003). The current study also reports a reliability of .79 for the instrument.

### Procedure

Ethical clearance was granted (ID-UCCIRB/CES/2022/30). To collect the data, permission was sought from household heads and the participants and the purpose of the study well explained to them.

A consent form was included in the IPAQ-Short form instrument. Participants read the form and consented by signing the consent form. Participants who could not read nor write, consent was orally explained to them and those who agreed to take part in the study thumb printed the consent form to give their approval. For inclusion criteria, only older adults who could walk and raise their upper limbs were considered for this study.

Participants were also assured of confidentiality of their information. Also, for purposes of anonymity, names of participants names were not taken.

First aiders were on standby during the fitness test to take care of any eventualities. Data was collected from participants at their homes. Physical function capacity for each participant was assessed through the five physical functional test batteries. To assess physical activity levels of each participant, IPAQ elderly short form was used and each participant indicated the number of days per week they walked, did moderate and vigorous activities or the amount of time they spent doing such activities. The 2-minute step-up test was used to assess cardio-respiratory endurance. A 2-minute test is conducted by marking the equivalent midpoint between the iliac crest and knee cap on a wall, and then, under instruction from the tester, the participant quickly steps up (lifts) each knee to the level of the mark on the wall. Each participant’s total number of step-ups within two minutes is noted. The data were collected between August and December, 2022 between 8 AM and 4 PM during weekends when the participants were likely to be at home.

### Data Analysis

Statistical Package for Social Science, IBM, version 25 for windows was used to manage the data which went through a thorough screening process before statistical analysis. A test for normality and linearity were also carried out. Demographic data of gender was analysed using frequency and percentages. Physical function capacity and cardio-respiratory function levels of older adults were analysed using frequency and percentages, because it helps to describe the portion of respondents measured on a categorical scale (Mishra et al., 2019). PFC levels were categorised as poor and improved. CRE was also categorized as low and recommended. Finally, multiple linear regression using PROCESS macro was computed to determine the mediating effect of physical activity on cardio-respiratory function and physical function capacity of the older adults. All the variables were measured on a continuous scale and the results were reported on the direct effect, indirect effect and total effect.

## Results

Frequency and percentages were used to display the cardio-respiratory function and physical function capacity levels of older adults in Navrongo. The level of cardio-respiratory function below the mean score of 61 steps-ups are considered poor and 61 and above step-ups are considered high and that such aged is meeting recommended cardio-respiratory function levels. Also, physical function capacity levels below the mean score of 90 are considered poor and above the mean are considered improved physical function capacity (see Table for details).

Demographic characteristics of participants was obtained using frequency and percentages

Table 1 above showed that, there were 565 males representing 56.6% and 433 females representing 43.4%.

**Table 1:**
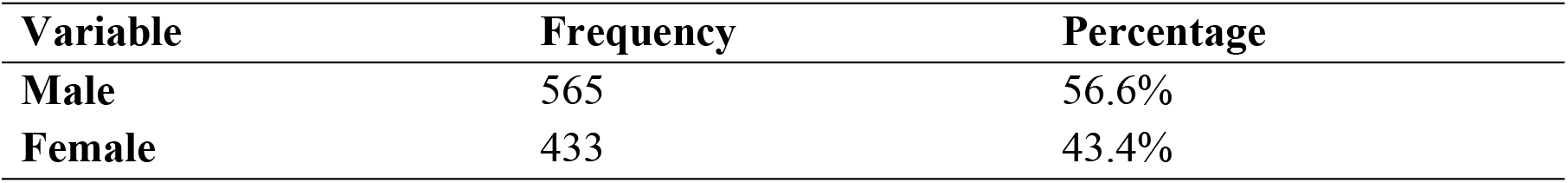
Data showing number of males and females in the study.

| Variable | Frequency | Percentage |
| --- | --- | --- |
| Male | 565 | 56.6% |
| Female | 433 | 43.4% |

To determine the mediating effect of physical activity participation on the relationship between cardio-respiratory function and physical function capacity levels, a simple mediation analysis was conducted using the regression model. The mediation model used PROCESS macro for SPSS (2022) model, at an alpha of .05. The mediation model was generally statistically significant, F (1, 996) = 1858.5, p < .05, and that cardio-respiratory function significantly influenced physical activity participation (β = .847, t = 2.089 p < .05), while, physical activity also significantly predicted physical function capacity of the older adults (β = .004, t = 2.238, p < .05). The direct effect of cardio-respiratory function on physical function capacity was also statistically significant (β = .867, t = 42.954, p < .05) and the indirect effect of CRF on PFC (β = .0030, t = 1.579 p < .05). Also, the total effect was also significant (β = .872, t = 43.110, p < .05). Hence, PA partially mediates the relationship between CRF and PFC of older adults in Navrongo (see Table 2 for details).

**Table 2:** Results showing CRF and PFC levels of Older Adults in Navrongo.

| CRF Range | Description | Frequency | Percentage (%) |
| --- | --- | --- | --- |
| (< 60 step-up/2 minutes) | Poor | 533 | 53.4% |
| (> 60 step-up/2 minutes) | Recommended | 465 | 46.6% |
| PFC Range |  |  |  |
| (< 90) | Poor | 538 | 53.9% |
| (> 90) | Improved | 460 | 46.1% |

**Table 3:** Mediation Analysis of the Effect of PA on the Relationship Between CRF and PFC of Older Adults in Navrongo.

| Variable | Coefficient | Standard Error | 95% Confidence Interval<br>Lower Bound'–Upper Bound' |  | t-value | p-value |
| --- | --- | --- | --- | --- | --- | --- |
| Direct effect | .867 | .0202 | .829 | -. 908 | 42.954 | <.05 |
| Indirect effect | .0030 | .0018 | .000 | - .007 | 1.579 | <.05 |
| Total effect | .872 | .0202 | .832 | -. 911 | 43.110 | <.05 |

**Figure 1:**
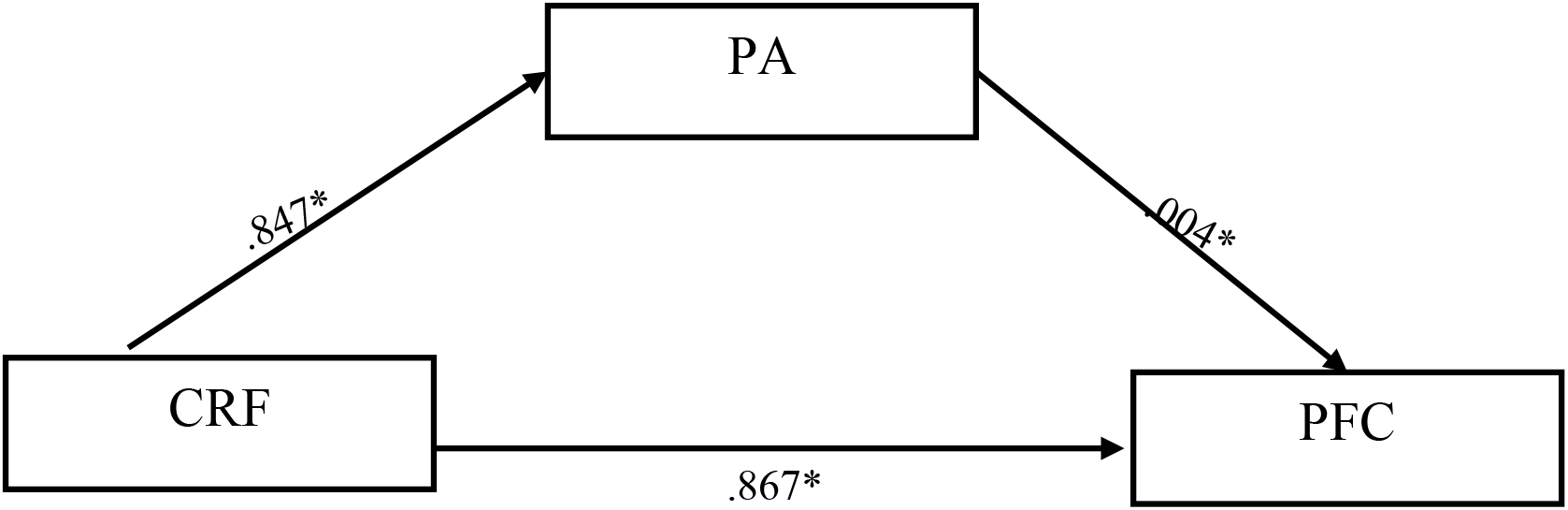
Mediation Effect of PA on the Relationship Between CRF and PFC of Older Adults in Navrongo. Keys: CRF = cardio-respiratory function; PA = physical activity; PFC = physical function capacity

## Discussions

The findings suggest that older adults in Navrongo had poor cardio-respiratory endurance levels as well as poor physical function capacity. Moreover, physical activity was found to partially mediate the relationship between cardio-respiratory endurance and physical function capacity among these older adults.

The indication of poor cardio-respiratory endurance levels of these older adults calls for a concern because it highlights a potential risk including age-related decline in physical function, independence, and overall quality of life. The fact that 53.4% of older adults in this study recorded low cardio-respiratory endurance levels is particularly alarming, as low cardio-respiratory function has been associated with a range of negative health outcomes among older adults, including cardiovascular disease, metabolic disorders, and cognitive decline (Myers, Kokkinos, Arena, & LaMonte, 2021). Such a low cardio-respiratory function levels may be attributed to several factors, including a sedentary lifestyle, poor nutrition, and associated chronic health conditions (Després, 2016). Additionally, older adults in Navrongo are more likely to lead sedentary lifestyles due to cultural factors, lack of access to physical activity resources, or limited knowledge about the benefits of exercise. According to Romero-Gómez, Zelber-Sagi, and Trenell (2017), poor nutrition, including inadequate caloric intake or nutrient deficiencies contribute to low cardio-respiratory endurance levels, because nutrition plays a critical role in supporting physical activity and exercise for health.

Chronic health conditions, such as CVDs, respiratory and metabolic disorders may further contribute to low cardio-respiratory function levels in older adults (Wang, & Ren, 2018). Furthermore, these conditions can impair oxygen uptake and utilization, making it more difficult for older adults to engage in meaningful physical activity and exercise. Moreover, chronic health conditions can lead to muscle wasting and decreased muscle strength, which further exacerbate declines in cardio-respiratory function of older adults. Thus, interventions aimed at improving cardio-respiratory function levels in older adults in Navrongo should focus on addressing these underlying risk factors. This may involve implementing community-based exercise programmes, promoting healthy eating habits, and providing access to healthcare services for the management of chronic health conditions (Suarez-Balcazar et. al., 2018). Additionally, educational campaigns aimed at increasing awareness about the benefits of exercise and physical activity in older adults may help to increase participation in these activities (McPhee et al., 2016). By promoting regular exercise and physical activity options, improving nutrition, and appropriately managing chronic health conditions, cardio-respiratory function may improve to promote better health outcomes among the older adults (Fletcher, Landolfo, Niebauer, Ozemek, Arena, & Lavie, 2018).

The finding again indicates that more than half (53.9%) of older adults in Navrongo recorded poor physical function capacity. Poor physical function capacity is a strong predictor of disability, falls, hospitalization, and mortality in older adults, and can significantly reduce their quality of life. The fact that such a high percentage of older adults in this study recorded poor physical function capacity highlights the need for evidenced-based interventions aimed at improving physical function in this population (Shimizu, Umemura, Matsunaga, & Hirai, 2018). Poor physical function capacity can be attributed to age-related declines in muscle strength, endurance, and flexibility, as well as chronic health conditions, because of environmental factors such as inadequate exercise facilities. Fragala et al. (2019) are of the view that as people age, they experience decline in muscle mass and function, which contribute to reduce mobility, balance, and overall physical function. According to Armstrong and Vogiatzis (2019), many age-related chronic health can also impair physical function by limiting oxygen uptake and utilization, reducing muscle strength, and causing fatigue.

Furthermore, environmental factors such as lack of physical activity facility including space further exacerbate sedentariness that in turn increase poor physical function capacity in these older adults (Lord & Close, 2018). For instance, inadequate healthcare services can result in the underdiagnose and under treatment of chronic health conditions, while limited access to physical activity resources make it difficult for older adults to engage in regular exercise (Armstrong & Vogiatzis, 2019). Thus, initiatives focused on improving physical function capacity in these older adults should aim at addressing these underlying risk factors (Anderson-Hanley et al., 2018). Additionally, there may be the need to implement community-based exercise programmes that focus on improving muscle strength, endurance, and flexibility, promoting healthy eating habits, and providing access to healthcare services for the management of chronic health conditions. Moreover, providing older adults with access to safe and supportive environments that promote physical activity can help to improve physical function capacity of these older adults. Generally, by addressing the underlying risk factors associated with poor physical function capacity, better health outcomes and improve the overall quality of life in this population are possible (Perpiñá-Galvañ et al., 2019).

The findings further support the hypothesis that physical activity participation mediates the relationship between cardio-respiratory endurance and physical function capacity levels in the older adults. The current finding demonstrate a significant relationship between cardio-respiratory function and physical function capacity, just as previous research (Harber et al., 2017). However, the novel contribution of this study is that it highlights the mediating role of physical activity, thus, reducing rate at which poor cardio-respiratory function compromise physical function capacity among these older adults (Tomás, Galán-Mercant, Carnero, & Fernandes, 2018). Thus, the more the older adults engage in physical activities including exercise, the more likely it is that their cardio-respiratory function will improve, which in turn promotes their physical function capacity.

We further found that cardio-respiratory function levels play important role in determining the level of physical activity participation (Edholm, Nilsson, & Kadi, 2019), which in turn impacts physical function in older adults (Laddu, Paluch, & LaMonte, 2021). This reinforces the importance of maintaining high levels of cardio-respiratory function via regular exercise. The direct effect of cardio-respiratory function on physical function capacity was also highlight the importance of cardio-respiratory function as a predictor of physical function in these older adults. This finding is consistent with previous research that demonstrated the importance of cardio-respiratory function in maintaining physical function capacity and independence in older adults (O’Neill, & Forman, 2020).

Overall, the results of the current study provide important insights into the relationship between cardio-respiratory function, physical activity participation, and physical function ability in older adults residing in Navrongo. The findings suggest that interventions aimed at improving physical function in older adults need to focus on promoting regular exercise, particularly in individuals with low levels of cardio-respiratory function (Izquierdo et al., 2021). Moreover, the results emphasize the importance of maintaining high levels of cardio-respiratory function through regular exercise and activity participation that promote physical function capacity and independence among these older adults.

### Limitations

Older persons’ physical functioning capacity may also be impacted by other factors like food, health issues, and drug use. The link between body weight, cardiovascular endurance, and physical functioning may therefore, be complicated by these uncontrolled variables. In an effort to study the mediating effect of physical activity on the relationship between cardio-respiratory function and physical function capacity of older persons, several of these confounding variables must therefore, be controlled.

### Practical Implications

The mediation model suggests a substantial correlation between older adults’ cardio-respiratory function, physical activity, and physical function capacity of these older adults. Accordingly, cardio-respiratory function positively affects physical function capacity both directly and indirectly through physical activity. Thus, raising cardio-respiratory function by consistent exercise can enhance older adults’ physical functional abilities.

The results also highlight the importance of considering both cardio-respiratory function and physical activity when designing interventions aimed at improving physical function capacity levels of these older adults. Thus, interventions that focus on one factor may not be as effective as interventions that address both or multiple factors. Overall, the mediation model provides important insights into the complex relationship between cardio-respiratory function, physical activity participation, and physical function capacity among these older adults. Such understanding should inform the development of effective interventions to improve physical function capacity among older adult population.

## Conclusions

Older adults in Navrongo recorded poor cardio-respiratory function levels as well as poor physical function and physical activity levels. Moreover, physical activity participation was found to partially mediate the relationship between cardio-respiratory endurance and physical function capacity of the older adults. These findings are an indication that these older adults are susceptible to heart-related diseases that might compromise or deteriorate their physical function capacity, thus creating poor quality of life among them. Therefore, an immediate evidence-based exercise intervention is required to attenuate the health consequences associated with poor cardio-respiratory function and poor physical function capacity among the older adults of Navrongo. These older adults are therefore recommended to engage in aerobic endurance activities as well as balance and strength training. Besides, creating an exercise-friendly community with adequate security would encourage exercise behavior among these older adults.

## Data Availability

The datasets generated and/or analysed during the current study are available in the Open Science Framework, and it is here: https://osf.io/zvdb2 data availability is subject to request from OSF and and approval by the corresponding author

## Declarations

### Ethics approval and consent to participate

Ethical clearance and approval were sought (ID-UCCIRB/CES/2022/30) for this study. all participants who consented to take part in this study duly signed or thumb printed a consent form provided to them.

### Consent for Publication

Informed consent was obtained from all subjects involved in the study. Participants and the heads of households were duly informed and they gave consent and approval that results and findings from this study could be published.

### Data Availability Statement

The datasets generated and/or analysed during the current study are available in the Open Science Framework, and it is here: https://osf.io/zvdb2

### Author Contributions

Conceptualization and study design: EAA and EWA. Methodology: EAA, EWA, EOS, and MS, Data collection: EAA, EWA, EOS, DA, and SRS, Data analysis: EAA and EWA, Writing - initial manuscript: EAA, Writing - final manuscript: EAA and EWA (with critical feedback provided by EOS, MS, and SRS and DA, Editing and review). All authors edited and considerably reviewed the manuscript, proofread for intellectual content and consented to its publication.

## Acknowledgements

The authors thank the people of Navrongo for their cooperation.

